# Subjective Cognitive Decline is a Better Marker for Future Cognitive Decline in Females than in Males

**DOI:** 10.1101/2022.08.18.22278960

**Authors:** Michael D. Oliver, Cassandra Morrison, Farooq Kamal, Jillian Graham, Mahsa Dadar

## Abstract

**Background:** The identification of biomarkers and other mechanisms for early detection of Alzheimer’s disease is critical to the development and further advancement of therapies and interventions targeted at managing symptoms and tracking the pathophysiology of disease. The endorsement of subjective cognitive decline (SCD) has emerged as a potential indicator of early change in cognitive status that may be predictive of future impairment at a time when measurable declines in neuropsychological performance cannot be detected. While there are numerous findings revealing sex differences in the prevalence of Alzheimer’s disease, there is a paucity of research examining sex differences in SCD. Therefore, the goal of this project is to determine if the relationship between the endorsement of SCD and future cognitive changes differ as a function of biological sex.

**Methods:** A sample of 3019 male and female healthy older adults (2188 without SCD, 831 with SCD), with a mean follow-up time of 5.7 years, were included from the Rush Alzheimer’s Disease Center Research Sharing Hub. Linear regressions were performed to determine group differences in baseline cognitive scores, while linear mixed effects models were computed to determine group differences in the rate of cognitive change over time.

**Results:** Individuals endorsing SCD had significantly lower baseline cognitive scores and increased rates of decline in all cognitive domains compared to those without SCD. Males exhibited significantly lower scores in baseline performance in global cognition, episodic memory, semantic memory, and perceptual speed regardless of SCD classification. Females with SCD were found to decline at significantly faster rates than both males with SCD and males and females without SCD in all cognitive domains over 15-year follow-up.

**Conclusions:** SCD is related to lower baseline cognitive performance and faster cognitive decline compared to those who do not endorse SCD. Females with SCD have the fastest rate of decline suggesting that SCD may be more predictive of future decline in females than in males. Therapeutic interventions targeting SCD in females may aid in the mitigation of sex disparities in AD prevalence.

## Introduction

Alzheimer’s disease (AD) is a neurodegenerative disorder characterized by the pathological aggregation of the proteins amyloid-β (Aβ) and tau in the brain (Selkoe, 1991). As Aβ plaques and neurofibrillary tau tangles form, communication between neurons is disrupted leading to atrophy, and ultimately functional impairment affecting multiple cognition domains (e.g., memory, visuospatial ability, language, and attention). Although debilitative functional changes occur with the progression of disease, it has been suggested that the pathophysiology of AD begins nearly 20 years prior to the clinical presentation of symptoms (Sperling et al., 2011; Younes et al., 2019). Therefore, it has become critical to target AD-related biomarkers early as they may be reflective of future decline. Advancements in clinical trial research have resulted in the US Food and Drug Administration’s (FDA) recent approval of Aducanumab (Aduhelm) for AD treatment. However, due to its lack of effectiveness at improving cognitive functioning, conflicting trial results, and potential harm caused by the drug (Knopman, Jones, & Greicius, 2021; Mahase, 2021; Tampi, Forester, & Agronin, 2021), agencies such as the European Medicines Agency (EMA) have refused to market this medication as a treatment for AD. As such, to date, there is still no uniform drug or treatment available to slow the progression of disease or reverse the disease process. For this reason, it has become increasingly important for research targeting mechanisms of early detection to help with disease prevention and to combat the deleterious effects of AD.

Research has suggested that subjective cognitive decline (SCD), or the self-reported experience of subtle changes in cognitive functioning without any measurable changes in neuropsychological test performance (Jessen et al., 2014), may be a preclinical marker of AD (Ávila-Villanueva & Fernández-Blázquez, 2017). For example, individuals who endorse SCD have an increased risk of developing AD compared to the general population (Rabin, Smart, & Amariglio, 2017; Ávila-Villanueva & Fernández-Blázquez, 2017; Slot et al., 2019). Typically reported as increased confusion or memory loss, the prevalence of SCD among adults aged 60 and older is around 25% (Röhr et al., 2020). Therefore, SCD may prove to be an effective target for early intervention. Existing intervention studies targeting AD risk factors have demonstrated success at reducing cognitive decline and dementia progression (Ngandu et al., 2015); however, these interventions are often introduced after the AD-related cognitive decline has already begun. SCD has been suggested as one of the earliest clinical indicators of AD prior to measurable cognitive decline, and its cognitive correlates align with the earliest pathological changes in AD. Therefore, explorations into SCD may allow for the improvement of early detection techniques at a critical time window prior to more pronounced atrophy and objective clinical symptoms as a consequence of disease progression.

Sex differences have also been observed in SCD; albeit findings yield inconsistent results. For example, it has been revealed that SCD in females is more strongly associated with future dementia diagnoses than in males (Heser et al., 2019). Conversely, another study has observed that SCD in males is associated with worse performance on a measure of global cognition (Alzheimer’s Disease Assessment Scale-13) compared to females (Wang et al., 2018). While the former suggests that SCD is associated with clinical progression in females more strongly than males, the latter indicates that SCD is associated with increased cognitive decline in males compared to females. Taken together, findings from these studies reveal that biological sex plays an integral role in SCD; however, further research is needed to better understand the interaction between sex and SCD. Despite evidence indicating biological sex is independently associated with AD prevalence (Cummings & Cole, 2002; Niu, Álvarez-Álvarez, Guillén-Grima, & Aguinaga-Ontoso, 2017), cortical atrophy (Filon et al., 2016; Koran, Wagener, & Hohman, 2017), and clinical progression (Fisher Bennett, & Dong, 2018; Zhu, Montagne, & Zhao, 2021), the relationship between sex, cognition, and SCD classification remains relatively unexplored.

It is critical to investigate whether cognitive decline observed in people with SCD differs as a function of biological sex. Such an exploration may result in a better understanding of whether females endorsing SCD have different cognitive trajectories subjecting them to greater decline and higher prevalence of AD compared to males. To examine this relationship, we investigated sex differences in cognition in a sample of female and male healthy older adults with and without SCD. The importance of examining sex differences in both groups is to ensure that the change over time is specific to those with SCD and not simply what occurs in healthy “normal” aging in this sample. This design allows us to determine whether SCD is predictive of future cognitive decline, and if this association differs by biological sex.

## Methods

### Participants

Data used in preparation of this article were obtained from the RADC Research Resource Sharing Hub (www.radc.rush.edu). Participants provided informed written consent to participate in one of three cohort studies on aging and dementia: 1) Minority Aging Research Study (Barnes et al., 2012), 2) Rush Alzheimer’s Disease Center Clinical Core (Schneider et al., 2009), or 3) the Rush Memory and Aging Project (Bennett et al., 2018).

Participant inclusion criteria for this specific study were as follows: 1) cognitively normal/healthy (NC/SCD−) status at their baseline visit (e.g., no mild cognitive impairment, MCI), 2) no report of stroke, 3) had completed at least two cognitive assessments, 4) completed the questionnaire assessing memory complaints, and 5) at least 55 years of age at baseline. Our two samples included a total of 3019 healthy older adult participants with a mean follow-up time of 5.7 years (with a total of 24689 follow-ups available for analysis, hereafter referred to as time from baseline). The healthy control (NC, no SCD/SCD−) sample (N=2188) had 528 or 24% of the sample as males. Similarly, the SCD sample (N=831) contained 196 males, or 24% of the sample as males.

Consistent with previous work investigating memory concerns in the RUSH cohort, subjective cognitive decline was defined based on two questions examining memory complaints (Barnes et al., 2006; Arvanitakis et al., 2018; Hill et al., 2020). Participants were asked, “About how often do you have trouble remembering things?” and “Compared to 10 years ago, would you say that your memory is much worse, a little worse, the same, a little better, or much better?” Both questions were scored using a scale of 1 to 5 with 5 being often/worse and 1 being never/much better. If the participants scored 8-10 on these two questions they were classified as having memory complaints; reported as subjective cognitive decline (SCD+) in this paper.

### Cognitive Assessment

All participants were administered a battery of neuropsychological tests including 19 tests selected to assess a range of five cognitive domains, and a measure of overall global cognitive function (Wilson et al., 2002; Barnes et al., 2016). There were seven tests of episodic memory (immediate and delayed recall of Story A of the Wechsler Memory Scale-Revised; immediate and delayed recall of the East Boston Story; Word List Memory, Recall and Recognition), three tests of semantic memory (Verbal Fluency; Boston Naming; Reading Test), three tests of working memory (Digit Span forward and backward; Digit Ordering), four tests of perceptual speed (Symbol Digit Modalities Test; Number Comparison; two indices from a modified version of the Stroop Test), and two tests of visuospatial ability (Line Orientation; Progressive Matrices). Composite measures of each domain were used in analyses, as well as a global composite of all tests. To create each composite score, individual tests were converted to z-scores, using the mean and standard deviation from the combined cohort at baseline, and z-scores for the relevant tests were averaged. More information for the specific tests used for each category can be obtained from https://www.radc.rush.edu/.

### Statistical Analysis

Analyses were performed using ‘R’ software version 4.0.5. Independent sample t-tests were completed on age and education. Multiple comparisons were corrected for using Bonferroni correction. Differences in baseline cognitive scores between male and females with and without SCD were examined using linear regressions. Rate of change in cognition between males and females were investigated using linear mixed effects models. These models examined the association between each cognitive domain (i.e., global, episodic memory, semantic memory, perceptual speed, working memory, and visuospatial abilities), SCD classification (i.e., SCD+ and SCD−), and Sex (i.e., Male and Female). All models were corrected for multiple comparisons using false discovery rate (FDR) (Benjamini & Hochberg, 1995); p-values were reported as raw values with significance, then determined by FDR correction.

To investigate the influence of sex on baseline cognitive scores by SCD classification, separate models were computed for SCD+ and SCD− using linear regressions with the categorical variable of interest of Sex (i.e., Male vs Female). The models also included years of Education and Age (at baseline, Age_bl) as covariates.

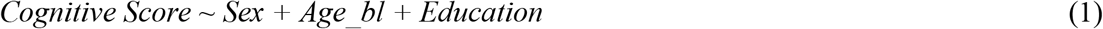

For the longitudinal analysis, the categorical variables of interest were Sex (i.e., Male vs Female), contrasting the males against the females, and SCD classification (i.e., SCD+ vs SCD−), contrasting SCD+ against SCD−. The models also included time from baseline, years of education, and age at baseline (*Age_bl*) as covariates. The interactions of interest were Sex:TimeFromBaseline, SCD:TimeFromBaseline, and Sex:SCD:TimeFromBaseline to examine if change over time differed between males and females within each group. Participant ID was included as a categorical random effect to account for repeated measures of the same participant.

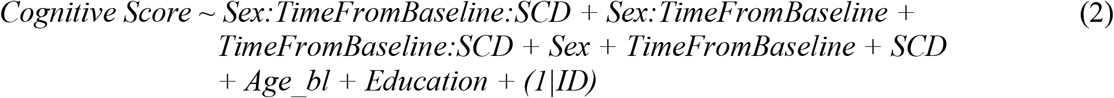

## Results

### Demographics and Baseline Cognitive Scores

In both SCD groups, males had higher education than females (SCD+: *t* = 4.30, *p*<.001; SCD−: *t* = 4.23, *p*<.001). Age did not significantly differ between SCD+ males and females (*t*=1.96) or SCD− males and females (*t*=1.55).

Figure 1a plots baseline cognitive scores for female and male SCD+ participants. Figure 1b plots baseline cognitive scores for female and male SCD− participants. Table 1 provides the outputs for the baseline linear regression models. For both SCD+ and SCD−, increased age was associated with lower cognitive scores at baseline in all cognitive domains (*t* belongs to [−5.75 – −12.51], *p*<.001) except visuospatial ability and working memory. On the other hand, increased education was associated with higher cognitive scores in all domains (*t* belongs to [10.22 – 25.13], *p*<.001). In addition, males had lower global cognition, episodic memory, and processing speed (SCD+: *t* belongs to [−2.47 – −3.83], *p*<.01; SCD−: *t* belongs to [−4.00 – −7.21], *p*<.001), but higher visuospatial scores (SCD+: *t*=5.35, *p*<.001; SCD−: *t*=7.41, *p*<.001) compared to females at baseline, regardless of SCD Classification. Further, semantic memory and working memory did not differ between males and females in either group.

**Table 1:** Linear regression outputs for baseline data

|  | Global Cognition | Episodic Memory | Semantic Memory | Perceptual Speed | Visuospatial Abilities | Working Memory |
| --- | --- | --- | --- | --- | --- | --- |
| SCD+ |  |  |  |  |  |  |
| Age at baseline | $t = -6.19$,<br>$p < .001$ | $t = -5.75$,<br>$p < .001$ | $t = -5.83$,<br>$p < .001$ | $t = -8.71$,<br>$p < .001$ | $t = 0.66$,<br>$p = .51$ | $t = 0.95$,<br>$p = 0.34$ |
| Male sex | $t = -2.46$,<br>$p = .014$ | $t = -3.33$,<br>$p < .001$ | $t = -1.33$,<br>$p = .19$ | $t = -3.83$,<br>$p < .001$ | $t = 5.35$,<br>$p < .001$ | $t = -1.71$,<br>$p = .09$ |
| Education | $t = 16.80$,<br>$p < .001$ | $t = 10.63$,<br>$p < .001$ | $t = 10.22$,<br>$p < .001$ | $t = 12.46$,<br>$p < .001$ | $t = 11.98$,<br>$p < .001$ | $t = 11.03$,<br>$p < .001$ |
| SCD- |  |  |  |  |  |  |
| Age at baseline | $t = -8.89$,<br>$p < .001$ | $t = -8.76$,<br>$p < .001$ | $t = -6.74$,<br>$p < .001$ | $t = -12.51$,<br>$p < .001$ | $t = -1.54$,<br>$p = 0.12$ | $t = 1.38$,<br>$p = .17$ |
| Male sex | $t = -4.00$,<br>$p < .001$ | $t = -7.21$,<br>$p < .001$ | $t = -1.97$,<br>$p = .049$ | $t = -5.33$,<br>$p < .001$ | $t = 7.41$,<br>$p < .001$ | $t = -0.22$,<br>$p = .83$ |
| Education | $t = 25.13$,<br>$p < .001$ | $t = 18.14$,<br>$p < .001$ | $t = 15.27$,<br>$p < .001$ | $t = 17.17$,<br>$p < .001$ | $t = 16.42$,<br>$p < .001$ | $t = 17.26$,<br>$p < .001$ |

Notes: Bolded values are results that remained significant after FDR correction.

**Figure 1:**
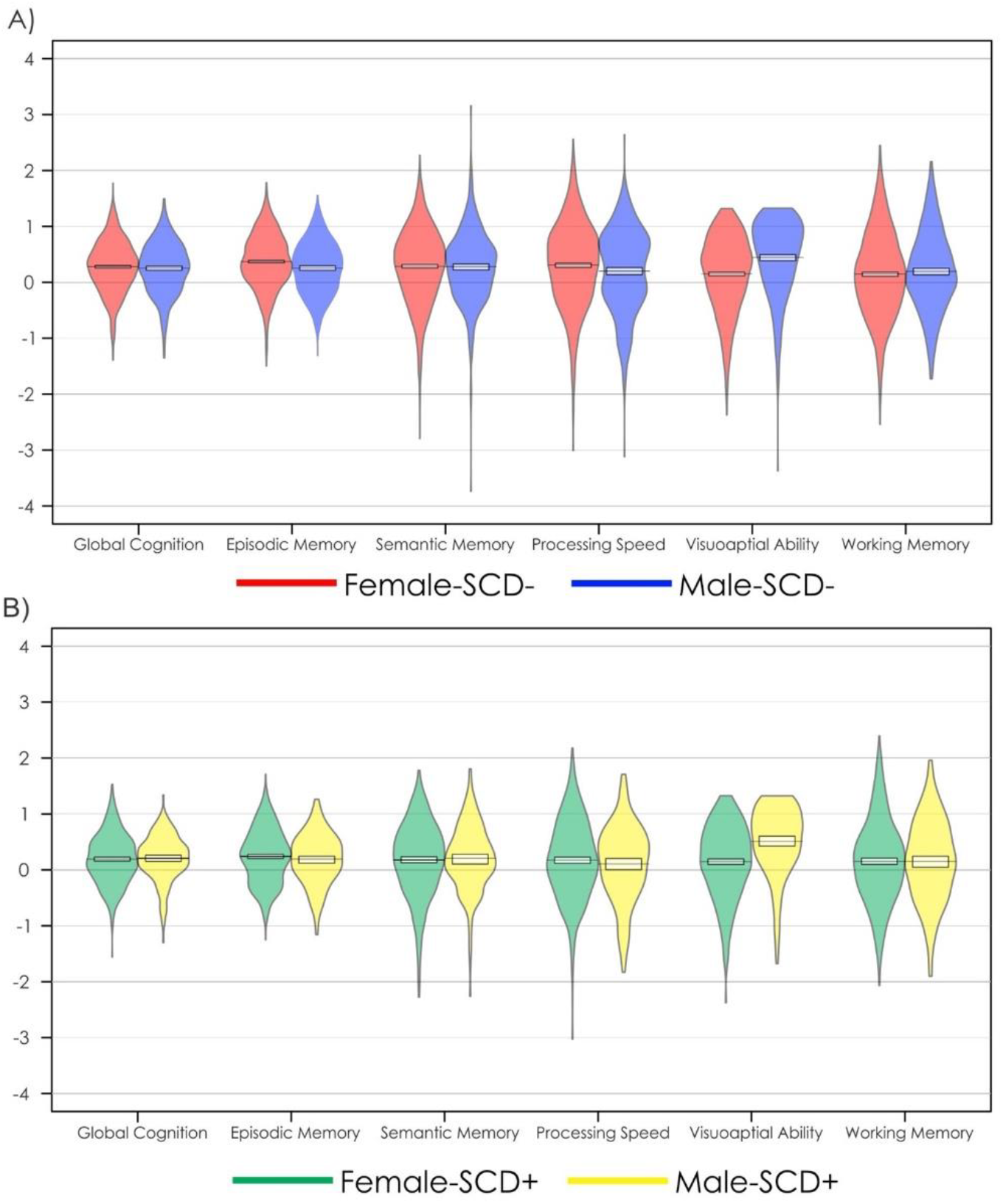
Baseline cognitive differences in each domain for females and males with and without subjective cognitive decline *Notes:* A) Baseline cognitive scores with mean and standard deviation for healthy older adults without subjective cognitive decline (SCD–). B) Baseline cognitive scores with mean and standard deviation for healthy older adults with subjective cognitive decline (SCD+).

### Cognitive Change

Figure 2 shows the mixed effects model predictions of cognitive scores over time for each cognitive domain by Sex and SCD Classification. Table 2 provides the estimates for the mixed effects model. For all cognitive domains, Time from baseline (*t* belongs to [−11.60 – −53.54], *p*<.001) and increased Age (*t* belongs to [−4.08 – −22.48], *p*<.001) were associated with lower cognitive performance. Increased Education was associated with increased scores in all cognitive domains (*t* belongs to [17.26 – 25.40], *p*<.001). All results remained significant after FDR correction.

**Table 2:** Linear mixed effects output for longitudinal data

|  | Global Cognition | Episodic Memory | Semantic Memory | Perceptual Speed | Visuospatial Abilities | Working Memory |
| --- | --- | --- | --- | --- | --- | --- |
| Age at Baseline | $t = -19.57$,<br>$p < .001$ | $t = -19.31$,<br>$p < .001$ | $t = -16.96$,<br>$p < .001$ | $t = -22.48$,<br>$p < .001$ | $t = -6.78$,<br>$p < .001$ | $t = -4.08$,<br>$p < .001$ |
| Male sex | $t = -3.95$,<br>$p < .001$ | $t = -6.90$,<br>$p < .001$ | $t = -2.31$,<br>$p = .02$ | $t = -5.64$,<br>$p < .001$ | $t = 8.39$,<br>$p < .001$ | $t = -0.10$,<br>$p = .92$ |
| Education | $t = 25.40$,<br>$p < .001$ | $t = 17.98$,<br>$p < .001$ | $t = 17.26$,<br>$p < .001$ | $t = 20.43$,<br>$p < .001$ | $t = 21.32$,<br>$p < .001$ | $t = 21.52$,<br>$p < .001$ |
| SCD Classification | $t = -1.65$,<br>$p = .097$ | $t = -2.53$,<br>$p = .011$ | $t = -1.87$,<br>$p = .061$ | $t = -2.12$,<br>$p = .035$ | $t = 0.87$,<br>$p = .39$ | $t = -1.00$,<br>$p = .31$ |
| TimeFromBaseline | $t = -35.82$,<br>$p < .001$ | $t = -19.02$,<br>$p < .001$ | $t = -34.50$,<br>$p < .001$ | $t = -53.54$,<br>$p < .001$ | $t = -11.60$,<br>$p < .001$ | $t = -17.43$,<br>$p < .001$ |
| Male sex: SCD Classification | $t = -0.43$,<br>$p = .67$ | $t = -0.43$,<br>$p = .66$ | $t = -0.43$,<br>$p = .67$ | $t = -0.72$,<br>$p = .47$ | $t = -0.36$,<br>$p = .71$ | $t = -1.17$,<br>$p = .24$ |
| SCD Classification: TimeFromBaseline | $t = -13.85$,<br>$p < .001$ | $t = -11.88$,<br>$p < .001$ | $t = -11.49$,<br>$p < .001$ | $t = -7.07$,<br>$p < .001$ | $t = -7.02$,<br>$p < .001$ | $t = -7.20$,<br>$p < .001$ |
| Male sex: TimeFromBaseline | $t = -1.59$,<br>$p = .11$ | $t = -0.25$,<br>$p = .80$ | $t = -1.78$,<br>$p = .07$ | $t = -0.37$,<br>$p = .71$ | $t = -1.02$,<br>$p = .30$ | $t = -3.72$,<br>$p < .001$ |
| Male sex: SCD Classification: TimeFromBaseline | $t = 4.97$,<br>$p < .001$ | $t = 3.56$,<br>$p < .001$ | $t = 3.22$,<br>$p = .001$ | $t = 2.96$,<br>$p = .003$ | $t = 4.00$,<br>$p < .001$ | $t = 2.84$,<br>$p = .004$ |

Notes: Bolded values are results that remained significant after FDR correction.

**Figure 2:**
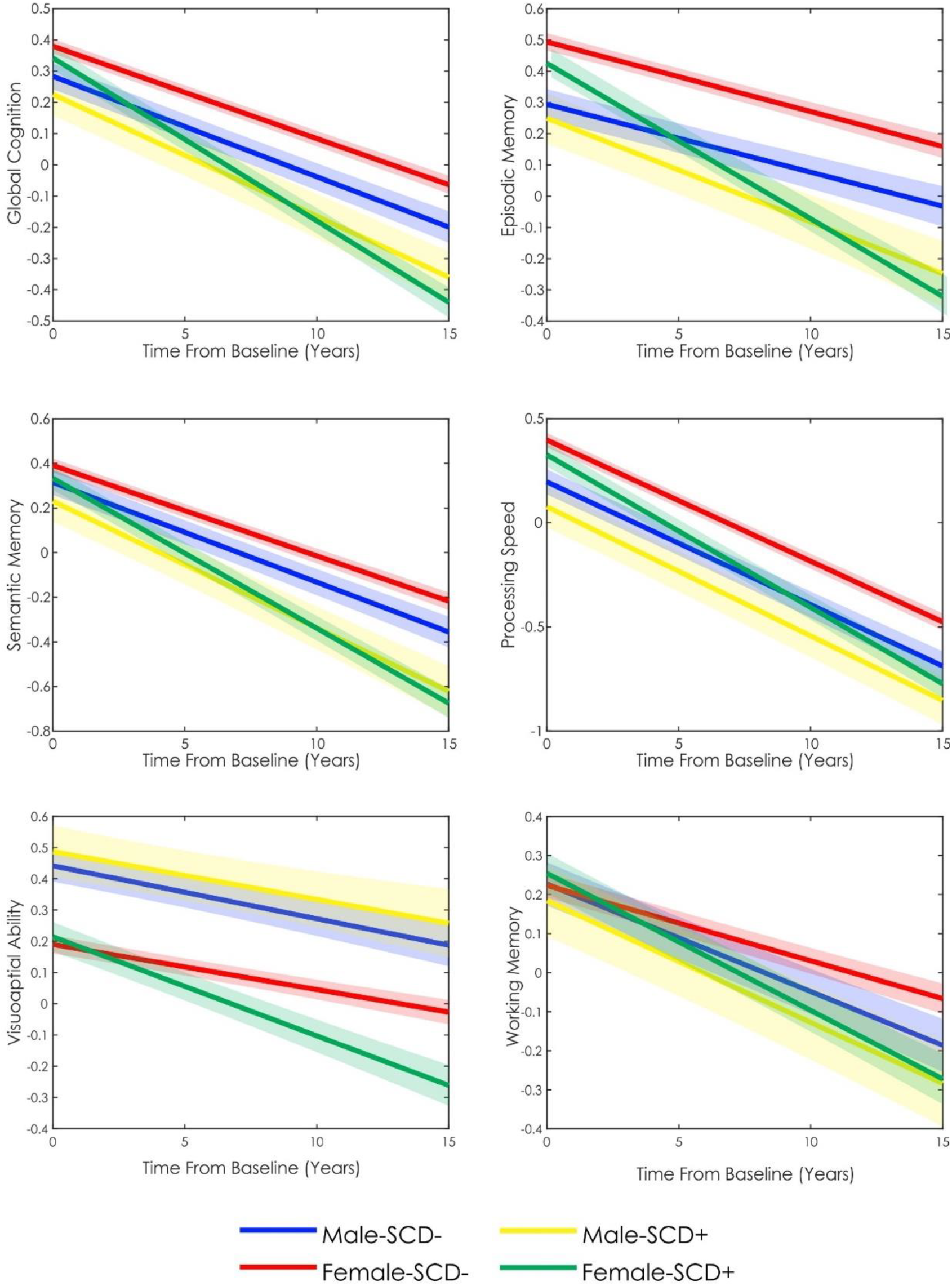
Longitudinal cognitive change over time in females and males with and without subjective cognitive decline *Notes:* SCD– = older adult without subjective cognitive decline. SCD+ = older adult with subjective cognitive decline

The main effect of SCD was significant for only episodic memory after FDR correction (*t* = −2.53, *p*=.011). The main effect of Male Sex was significant for all domains except working memory. Males exhibited lower overall performance in global cognition, episodic memory, semantic memory, and perceptual speed (*t* belongs to [−2.31 – −6.90], *p*<.05), but higher overall performance in visuospatial abilities (*t* = 8.39, *p*<.001). The Male by SCD Classification was not significant for any cognitive domain, and the Male by Time From Baseline year was only significant for working memory (*t* = −3.72, *p<*.001). The SCD Classification by Time From Baseline interaction was significant for all cognitive domains (*t* belongs to [−7.02 – −13.85], *p*<.001), indicating that people with SCD had increased rates of decline in all domains compared to SCD–. The three-way interaction between Male Sex, SCD Classification, and Time From Baseline was significant for all cognitive domains (*t* belongs to [2.84 – 4.97], *p*<.005). Taken together, these results suggest that SCD+ females decline at significantly faster rates than SCD− females in all cognitive domains (*t* belongs to [−7.02 – −13.85], *p*<.005), whereas SCD+ males decline at faster rates than SCD− males in global cognition, episodic memory, and semantic memory (*t* belongs to [−2.51 – 3.12], *p*<.01). Moreover, while SCD− males and females do not differ in terms of rate of cognitive decline in any cognitive domain except for working memory, where males exhibit a faster decline, SCD+ females decline at significantly faster rates than SCD+ males in all cognitive domains (Figure 2, Table 2).

## Discussion

Previous findings have suggested that cognitive functioning, including rate of cognitive decline, may differ between males and females (e.g., Levine et al., 2021). However, there is limited research examining sex differences in people who may be at the earliest stages of cognitive decline; those with SCD. This limited understanding of how sex may influence cognitive decline in SCD limits the ability to have targeted interventions and therapies to help prevent cognitive decline due to MCI or dementia. Therefore, the current study aimed to elucidate sex disparities in the trajectory of cognitive decline to aid in a better understanding of techniques for early detection and disease mitigation. In our sample of 3019 cognitively unimpaired older adults, the rate of change in cognitive performance varied between males and females in people with SCD. Specifically, males exhibited significantly lower baseline performance in global cognition, episodic memory, semantic memory, and perceptual speed, but higher performance in visuospatial abilities. The three-way interaction between Sex, SCD classification, and Time from baseline was also significant, revealing that SCD+ females decline at a significantly faster rate than SCD+ males in all cognitive domains. When examining the effect of SCD classification on overall cognitive performance collapsed across sex, a main effect of SCD classification revealed that SCD+ exhibited significantly lower performance in episodic memory compared to SCD−. Over time, SCD+ individuals, regardless of sex had a significantly faster rate of decline in all domains compared to SCD−. Our results reveal that 1) people with SCD have both lower baseline cognition and an increased rate of decline compared to people without SCD, and 2) SCD in females may be more predictive of future cognitive decline than in males; thus, contributing to sex disparities in cognitive function.

There is mounting evidence suggesting that sex differences exist regarding both normal cognition and dementia. For example, a recent cohort study of over 26,000 participants reported cognitively unimpaired females to have greater global cognition, executive function, and memory compared to males (Levine et al., 2021). Similarly, in our study, a main effect of sex was observed for all cognitive domains except working memory, demonstrating that regardless of SCD status, females tend to score higher on neuropsychological assessments in several domains compared to males, whereas males score higher in visuospatial ability then females. Levine and colleagues (2021) also observed that cognitively unimpaired females had an increased rate of decline compared to males. Additionally, several other studies have suggested that although females may score higher at baseline, they may be subject to faster cognitive decline compared to males (Irvine, Laws, Gale, & Kondel, 2012; Holland, Desikan, Dale, & McEvoy, 2013; Lin et al., 2015). This increased rate of decline in females may contribute to the sex disparities that exist in prevalence of AD (Beam et al., 2018; Liu, Li, Sun, & Hu, 2019). In the present study, females were not observed to have an increased rate of decline in the SCD− group compared to males. Rather, sex differences in cognitive decline were observed only in SCD+ group. SCD+ females experienced steeper declines in cognitive performance compared to SCD+ males, and SCD− males and females, in all domains. That is, although SCD− females are shown to consistently have the highest cognitive performance over time compared to SCD− males (except in visuospatial abilities), and SCD+ males and females, the introduction of SCD (+), negatively affects this relationship causing females to decline much more rapidly. Our findings are consistent with a similar study exploring sex differences in dementia incidence and prevalence as a function of SCD status reporting that SCD in females was more strongly associated with subsequent dementia compared to males (Heser et al., 2019).

Given that SCD (SCD+) has been linked to increased risk for MCI and AD (Jessen et al., 2020), those who endorse these subjective cognitive complaints are at greater risk for subsequent cognitive decline compared to those that do not. In the current study, both male and female SCD+ participants had increased rate of decline compared to SCD− participants, supporting the notion that SCD is indicative of future decline (Koppara et al., 2015; Jessen et al., 2020).

However, SCD+ males only had increased rates of decline in global cognition, episodic memory, and semantic memory compared to SCD− males, whereas SCD+ females decline at significantly faster rates than SCD− females in all cognitive domains. Furthermore, with the presence of sex differences in SCD+ (i.e., SCD+ females exhibiting increased rates of change in all cognitive domains compared to SCD+ males), our findings suggest that the relationship between SCD and future cognitive changes may be more predictive of cognitive decline in females compared to males. That is, females reporting SCD may be more likely to experience substantial cognitive changes compared to males with SCD. These findings suggest that waning cognitive abilities may have the potential to be captured early, particularly in females, with SCD+ individuals detecting subtle cognitive changes prior to objective testing. Previous studies have observed that females tend to self-report cognitive changes more than males (Martinez et al., 2021). Combining with our findings with the increased reports in females relative to males may be indicative of either greater changes, or better perception of cognitive changes in females. As such, the relationship between SCD and cognition may be stronger in females.

Findings from this study have important implications for interventions and therapies designed to target cognitive decline and dementia prevention. For example, risk factors such as midlife hypertension, midlife obesity, diabetes, physical inactivity, smoking, depression, and low education are all modifiable factors contributing to 1/3rd of all AD cases (Norton, Matthews, Barnes, Yaffe, & Brayne, 2014). However, several of these AD risk factors disproportionately affect females. For example, both lower educational attainment, as well as psychiatric disorders such as depression, are more prevalent in females (Huebschmann et al., 2019). Additionally, blood pressure is observed to be higher in males early in life, whereas females have a steeper increase in blood pressure that continues throughout the life compared to males (Ji et al., 2020). This prevalence of higher mid-life blood pressure in females is associated with a greater risk for the development of dementia compared to males (Blanken & Nation, 2020). Other factors specific to females such as preeclampsia, menopause, and hypertensive pregnancy disorders also have negative impacts on the cardiovascular system and cognition (Gannon et al., 2019; Miller et al., 2013). These risk factors, paired with explanations such as higher life expectancy (Hebert, Scherr, McCann, Beckett, & Evans, 2001), lower cognitive reserve, and faster rates of functional and structural deterioration (Laws, Irvine, & Gale, 2018) in females compared to males have all led previous literature to reveal female sex to be a significant risk factor for AD (Beam et al., 2018; Liu, Li, Sun, & Hu, 2019). The current study supplements the existing literature by revealing that in the earliest potential stage of the AD-trajectory prior to measurable cognitive decline (i.e., preclinical-AD or SCD), females also exhibit steeper declines in all cognitive domains over time compared to males. As such, directing therapies and interventions toward risk factors that have increased incidence in females may help reduce the prevalence of dementia in these individuals. Our findings suggest that SCD may be a critical indicator of subsequent cognitive decline in females, and therapeutic interventions may wish to target this population to better elucidate sex disparities in cognitive change over time.

One limitation of the current work is use of only two questions to determine SCD status. Previous research has shown that different questionnaires used to determine SCD status results in different cognitive trajectories and atrophy patterns (Morrison et al., 2022) as well as different patterns of white matter hyperintensity burden (Morrison et al., 2022b). Therefore, it is thus possible that the use of different questionnaires may target specific declines in males vs. females and improve the relationship between SCD and cognition in males. Future research should explore this relationship. Females also have increased risk factors that influence vascular components. The resulting pathological changes due to vascular damage, such as white matter hyperintensities which are known to be associated with cognitive decline and conversion to dementia (Dadar et al., 2019), may be higher in females with SCD. Future research should examine the association between sex and SCD status on atrophy and white matter hyperintensities.

## Conclusion

The current study compared cognitively unimpaired males and females with and without SCD to demonstrate that sex differences influence rate of change in cognitive performance over time. Our findings suggest that while those with SCD have lower baseline cognitive scores compared to those without SCD, SCD may be more predictive of future decline in females than in males. These findings have implications for clinical and research settings where future prediction of cognitive decline and conversion to dementia are examined. Furthermore, these findings should be considered when developing interventions to slow progression of cognitive decline. Targeting intervention techniques on risk factors known to more prevalent in females may help reduce the increased incidence of dementia in females, with the overall goal of lowering the rate of decline and conversion to dementia.

## Data Availability

All data produced are available online after a written request and application is made at https://www.radc.rush.edu/

## Acknowledgments

We want to acknowledge all the MARS, AA Core, and MAP participants. We are also grateful for the hard work from the staff and investigators at the Rush Alzheimer’s Disease Center. To obtain data from MARS, AA Core, and MAP for research use, please visit the RADC Research Resource Sharing Hub (www.radc.rush.edu).

